# Copy number variant detection with low-coverage whole-genome sequencing is a viable alternative to the traditional array-CGH

**DOI:** 10.1101/2020.09.07.20183665

**Authors:** Marcel Kucharík, Jaroslav Budiš, Michaela Hýblová, Gabriel Minárik, Tomáš Szemes

## Abstract

Copy number variations (CNVs) are a type of structural variants involving alterations in the number of copies of specific regions of DNA, which can either be deleted or duplicated. CNVs contribute substantially to normal population variability; however, abnormal CNVs cause numerous genetic disorders. Nowadays, several methods for CNV detection are used, from the conventional cytogenetic analysis through microarray-based methods (aCGH) to next-generation sequencing (NGS). We present GenomeScreen – NGS-based CNV detection method for lowcoverage whole-genome sequencing. We determined the theoretical limits of its accuracy and confirmed it with extensive in-silico study and real patient samples with known genotypes. Theoretically, at least 6M uniquely mapped reads are required to detect CNV with a length of 100 kilobases (kb) or more with high confidence (Z-score > 7). In practice, the in-silico analysis showed the requirement of at least 8M to obtain >99% accuracy (for 100 kb deviations). We compared GenomeScreen with one of the currently used aCGH methods in diagnostic laboratories, which has a 200 kb mean resolution. GenomeScreen and aCGH both detected 59 deviations, GenomeScreen furthermore detected 134 other (usually) smaller variations. The performance of the proposed GenemoScreen tool is comparable or superior to the aCGH regarding accuracy, turnaround time, and cost-effectiveness, presenting a reasonable benefit particularly in a prenatal diagnosis setting.

## 1. Introduction

Copy number variations (CNVs) are a phenomenon in which sections of the genome are repeated, and the number of repeats in the genome varies between individuals. CNVs contribute substantially to normal population variability. However, abnormal CNVs are a cause of numerous genetic disorders. Several methods for CNV analysis are used, from the conventional cytogenetic analysis through microarray-based approaches to next-generation sequencing (NGS) [1].

Array-based comparative genomic hybridization (aCGH) provides genome-wide coverage at a great resolution, even on the scale of tens of kilobases (10–25 kb) [2]. This fact promoted aCGH for a golden standard in CNVs detection for several years. Even though current microarrays offer flexibility in coverage across variable resolution formats, there are still some disadvantages to consider. In prenatal diagnosis from amniotic fluid, for example, usually micrograms of genomic DNA are needed to hybridize to an array. This can be accomplished either by time-consuming cultivation of up to two weeks or whole genome amplification which can introduce bias into the analysis. On the contrary, NGS utilizes as little as nanograms of DNA and thus it does not need additional amplification and sample contamination is less likely due to less material required. The transition from the proven microarray platform to NGS, often revealing something new and unexpected, seems to be very slow, however, the cost and time aspect is already unprecedented. Besides, while aCGH equipment serves one purpose only, commonly used NGS platforms are very versatile for many applications whether exome, genome, targeted panels, transcriptome, or episome sequencing. The whole-exome and targeted sequencing aims to reduce the sequencing cost but is limited to certain regions (protein-coding, or custom), where most known disease-causing mutations occur [3]. NGS provides a sensitive and accurate approach for the detection of the major types of genomic variations, including CNVs [4,5].

Recent years yielded a handful of CNV detection tools specifically for targeted and exome sequencing [6–12], however, these tools are not suitable for data from whole-genome low-coverage sequencing. The notable whole-genome CNV detection tools include: Wisecondor X [13] (successor of Wisecondor [14] tool), CNVkit[15], CNVnator [16], iCopyDav [17]. The partial comparison of some of these tools is in the publication of Wisecondor X [13].

We present GenomeScreen – a low-coverage whole-genome NGS-based CNV detection method and estimate its accuracy in theoretical and in-silico settings. This method is partially based on the previously published NIPT CNV detection method [18,19]. The main differences are the parameters of reported CNVs - in the NIPT setting, CNVs corresponding to more than 5% fetal fraction and at least 3Mb in size were reported, here we focus on full (non-mosaic) aberrations with much shorter length (100 kb and larger). Furthermore, we compare the sensitivity of GenomeScreen to the more conventional aCGH method on 106 laboratory-prepared clinical samples. The comparison of GenomeScreen and different CNV detection tools are beyond the scope of this article due to focus on comparison with the aCGH method itself.

## 2. Materials and Methods

### 2.1 Sample collection and processing

All patient samples were analyzed as a part of commercially available testing in cooperation with gynecologists, clinical geneticists, and genetic centers. All patients signed informed consent for participation in the research. Samples of chorionic villi, amniotic fluid, placenta, tissue, or peripheral blood were obtained from 106 patients in the clinical sample group and 789 in the training group. Peripheral blood was drawn in EDTA or STRECK tubes, inverted several times after collection, stored in a chilled environment (4–10 °C) for EDTA and at room temperature for STRECK tubes, and transported to the laboratory within 36 hours. DNA was extracted from 200 µl of whole blood or 700 µl of amniotic fluid using the QIAamp DNA Blood Mini Kit (Qiagen, Hilden, Germany) according to the manufacturer’s protocol and stored at −20°C until further analysis.

Genomic DNA from clinical samples was fragmented using 1U/µl dsDNA Shearase™ Plus (Zymo Research, Irvine, CA, USA) and incubated 23 min at 42°C to generate 100-500bp fragments. For adapter-ligated DNA library construction, a TruSeq Nano kit (Illumina, San Diego, CA, USA) with an in-house optimized protocol was used. Low coverage sequencing (0.3×) was performed on Illumina NextSeq 500/550 platform (Illumina, San Diego, CA, USA) with paired-end setting 2×35 using High Output Sequencing Kit v2.5. Library quantity and quality were measured by fluorometric assay on Qubit 2.0 (ds DNA HS Assay Kit, Life Technologies, Eugene, Oregon, USA). Fragment analysis was performed on 2100 Bioanalyzer (High Sensitivity DNA Kit, Agilent Technologies, Waldbronn, Germany). We targeted 5M uniquely mapped reads per sample; however, none of the analyses were excluded due to lower (or higher) read counts (more info in Supplementary material Table 1.).

### 2.2 Theoretical minimal read count estimation

Suppose we model sequencing as a random choice of reads from the whole (mappable) genome. Then we can theoretically deduce the number of needed uniquely mapped reads for a certain accuracy criterion. The random choice for a target region is described by the binomial distribution with mean *μ*=*np* and variance *σ*^2^=*np*(1−*p*). Here, *p* is the probability of choosing a read from the target region, and *n* is the number of reads sequenced. The probability *p* can be furthermore expressed as the ratio of the region length *l*_*c*_ to whole-genome length *l*_*g*_ (*p*=*l*_*c*_ / *l*_*g*_). When predicting a CNV, we need to have certain confidence traditionally determined by the Z-score (*Z*), defined as:

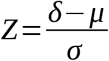

Here *δ* is the number of reads that we observe in the target region. We assume that the number of reads in the target region will be proportional to the number of present copies of gonosomes, i.e., either *δ* =*n* (*p* + *p* / 2) for duplication or *δ* =*n* (*p*− *p*/ 2) for deletion of the region on a single chromosome. If we solve for *Z*^2^ and substitute:

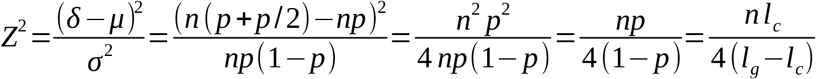

Then we can estimate the minimal number of reads (*n*) to be able to predict a variation with length *l*_*c*_ with the desired Z-score (*Z*):

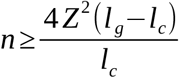

### 2.3 Variant identification

To identify variations, we performed the following pipeline:

1. Mapping and binning
  a. mapping reads using bowtie2 [20]
  b. binning reads into same-size 20 kb bins
  c. normalizing bin counts
2. Normalization (similar to one published previously by [21])
  a. LOESS-based GC correction [22]
  b. PCA normalization to remove higher-order population artifacts on autosomal chromosomes
  c. subtracting per-bin mean bin count to obtain data normalized around zero
3. Filtration of unusable bins
  a. unmappable or badly mappable regions (zero or low mean of bin count)
  b. repetitive regions or areas with some systematically increased mappability (high mean of bin count)
  c. highly variable regions (high variance of bin count)
4. Segment identification and reporting
  a. circular binary segmentation algorithm [23] to identify consistent segments of similar coverage
  b. assigning significance to segments based on the proportion of reads
  c. visualization of findings (Figure 1.)

**Figure 1.**
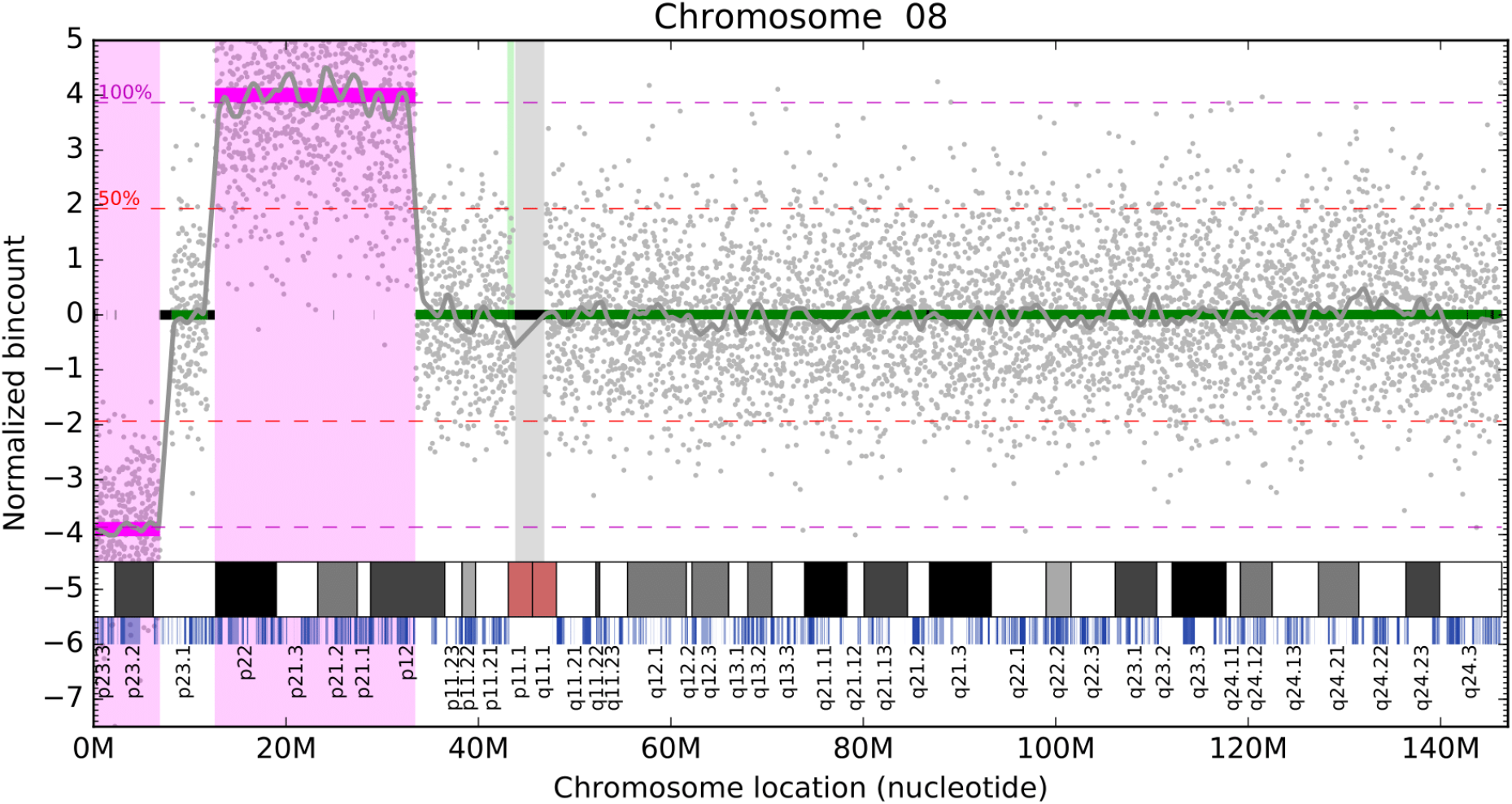
Visualization of detected deviations on chromosome 8. Chromosome location is on X-axis. Normalized bin count is on Y-axis. Green lines represent normal bin count segments (normalized around zero), magenta lines visualize aberrations (one deletion at the start of the chromosome, one duplication on p22-p12). Filtered bins are depicted as black bars on the zero line on Y-axis. The unmapped region around the centromere is visualized with a grey bar. Grey dots represent the normalized individual bin counts for each bin.

Scripts (Python 3.7) and data are available on the website https://github.com/marcelTBI/GenomeScreen.

#### 2.3.1 Mapping and binning

Firstly, the reads are mapped to a reference using Bowtie 2 [20] with *--very-sensitive* settings. We use hg19 reference in all applications, but other references can be used without changes to the algorithm. The reads are then filtered for map quality at least 40 and binned according to their starts to same-size 20 kb bins. All subsequent analyses are performed on the bin counts, and the algorithm does not use any other information about reads (for example, sequence). For training purposes, the bin counts corresponding to autosomal chromosomes for each sample are normalized to the same number of reads (i.e., each bin is divided so the sum of all bins on autosomal chromosomes would be the same for each sample). Furthermore, the same is done separately for chromosome X and chromosome Y. A consequence of separate normalization of sex chromosomes is that the current approach can detect only small sex chromosomal variations and not the whole sex chromosomal aneuploidies.

#### 2.3.2 Normalization

Normalization consists of three steps: firstly, a sample-wise LOESS-based GC correction is employed on the bin counts [22]. Afterward, the principal component analysis (PCA) normalization is used to remove higher-order population artifacts on autosomal chromosomes [21]. For training of the PCA, LOESS-corrected bin counts of 789 NIPT samples with female fetuses were converted to principal component space, and the first 15 principal components were stored. The bin count vector of a new sample is then transformed into principal component space defined by these first 15 components and transformed back to the bin space to obtain residuals that are then removed from the bin counts. The first principal components represent noise commonly seen in euploid samples, and their removal helps to normalize the data. Currently, the PCA normalization is done only on autosomal chromosomes due to the unavailability of enough male samples for training. In the future, the training of PCA on both male and female samples is likely to increase the prediction precision for sex chromosomes. Lastly, we subtract per-bin mean bin counts to obtain data normalized around zero. This last step is trained already on the PCA normalized bin counts (where available) and helps compensate for the mapping inequality between various genomic regions.

#### 2.3.3 Filtration of unusable bins

To further improve accuracy, we filter bins that have an unusual signature – either low mean (this signals bad mappability of the region), high mean (repetitive regions or regions with some systematic bias), or high variance (highly variable regions). Furthermore, the filtered regions were manually curated to reduce the scatter of filtered regions, mainly around centromeres and in sex chromosomes. The filtration leaves out around 15% of the genome, mainly due to the low mappability, especially in and around centromeres.

#### 2.3.4 Segment identification and reporting

After normalization and filtering, we have a signal (grey dots in Figure 1.) that needs to be segmented into the same level parts to be evaluated. For this purpose, we use the circular binary segmentation (CBS) algorithm implemented in the R package DNAcopy [23]. After segmentation, each segment is assigned a significance level based on its length and difference from zero. Since we know the mean bin counts, we can estimate the level for a complete deletion or duplication of one copy of a chromosome (magenta dashed lines in Figure 1.). We then differ between five color-coded levels of significance: magenta – at least 75%, at least 200 kb, red – at least 25%, at least 200 kb, orange – at least 25%, at least 40 kb, yellow – at least 12,5%, at least 40 kb, and green – all others (very short segments or segments around zero). The findings are then reported as a text file for further machine processing, and each chromosome is visualized (Figure 1.).

### 2.4 In-silico analysis

For *in-silico* analysis, we chose 83 samples without any aberration and with a read count of at least 10M. Firstly, the samples were downsampled to the studied read count (3M – 10M with the step of 1M). Then, for each of the tested variation lengths (20 kb – 200 kb with the step of 20 kb), 100 random variations on autosomal chromosomes were generated that do not overlap with the filtered regions (see Section 4.3.3). To create a sample with an artificial aberration, the bins corresponding to the generated random variation were multiplied accordingly (thus, the most time-consuming mapping step was performed only once per sample). Afterward, variant identification was performed without changes.

In total, we gradually created 664,000 artificial samples (100 variations * 83 samples * 10 variation lengths * 8 read counts) and performed variant identification on them to analyze the impact of read count and variant length. The results are displayed in Figure 3.

## 3. Results

### 3.1 Theoretical minimal read count

The theoretical minimum of reads for predicting a variation with length *l*_*c*_ with the desired Z-score (*Z*) is estimated as (see Section 4.2):

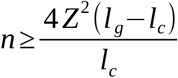

Standardly, a Z-score of 4 is used in the detection of whole chromosomal aneuploidies [24,25]; however, there are inherently more possible CNVs than whole chromosomal aneuploidies. Thus, the desired Z-score should be much higher in this instance to decrease the number of false positives. Moreover, in practice, the number of reads needed would be even larger due to the uncertainty of sequencing, mapping, and inherent biological biases [26,27]. The theoretical minimal read count estimation for different Z-scores can be seen in Figure 2.

**Figure 2.**
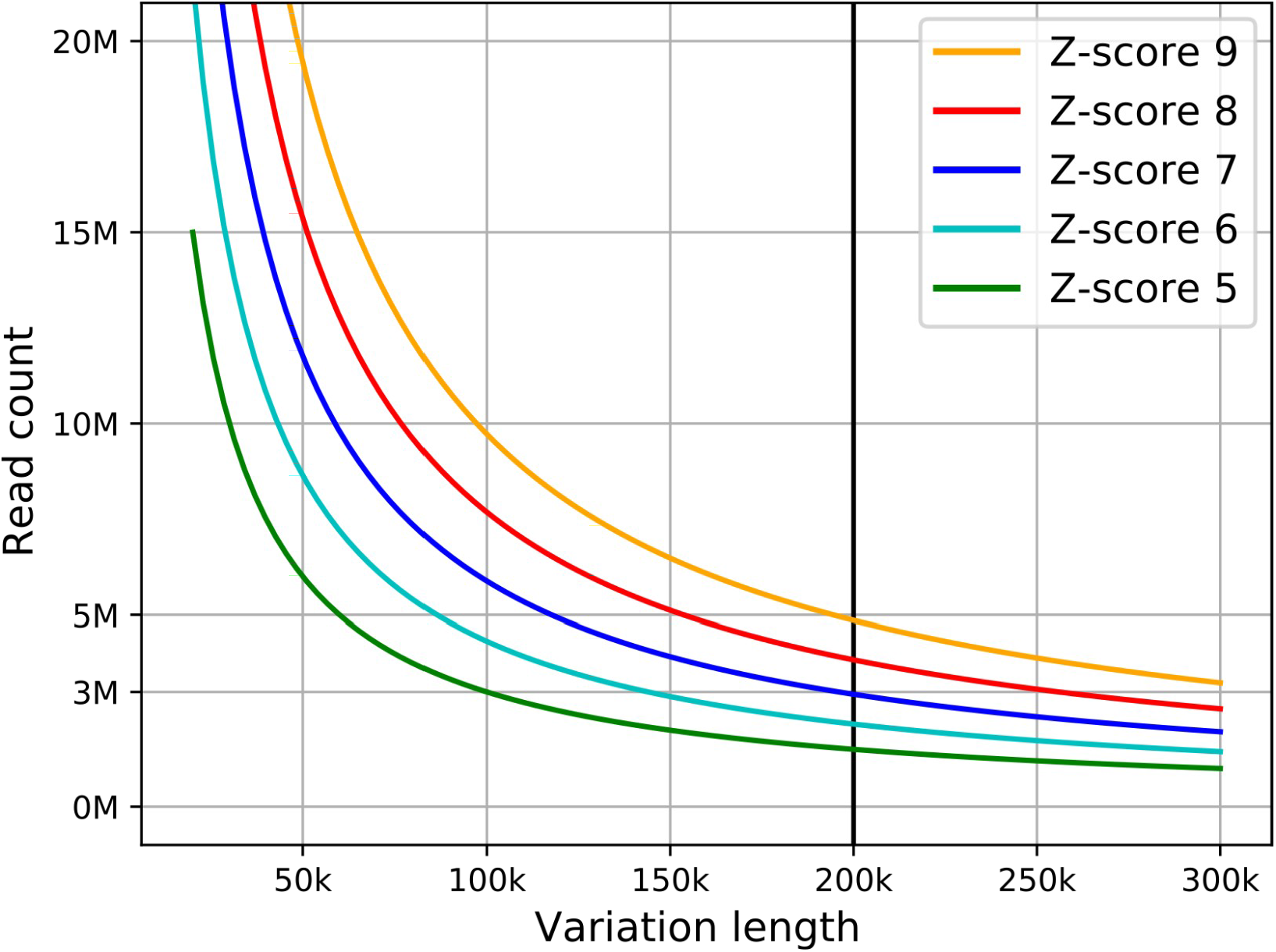
Theoretical minimal read count for successful estimation of CNV with specified variation length. Different lines represent different Z-score confidence levels.

### 3.2 Detection accuracy for variable CNV lengths and read count (in-silico)

To verify the theoretically estimated limitations, we first conducted a simulated *in-silico* experiment. Artificial samples with simulated CNV were created from healthy samples by multiplication of bins corresponding to the simulated region randomly on the genome. Only regions that did not span into filtered positions were kept for further analysis (about 85% of the genome). The details can be found in Section 4.3.

The *in-silico* analysis shows the influence of read count and CNV length for prediction accuracy (Figure 3.). Based on the findings, we recommend using a read count of at least 8M to achieve >99% prediction accuracy for variations with 100 kb and more. Thus we recommend following the line for a Z-score of 8 (red on Figure 2.) for estimation for different CNV lengths.

**Figure 3.**
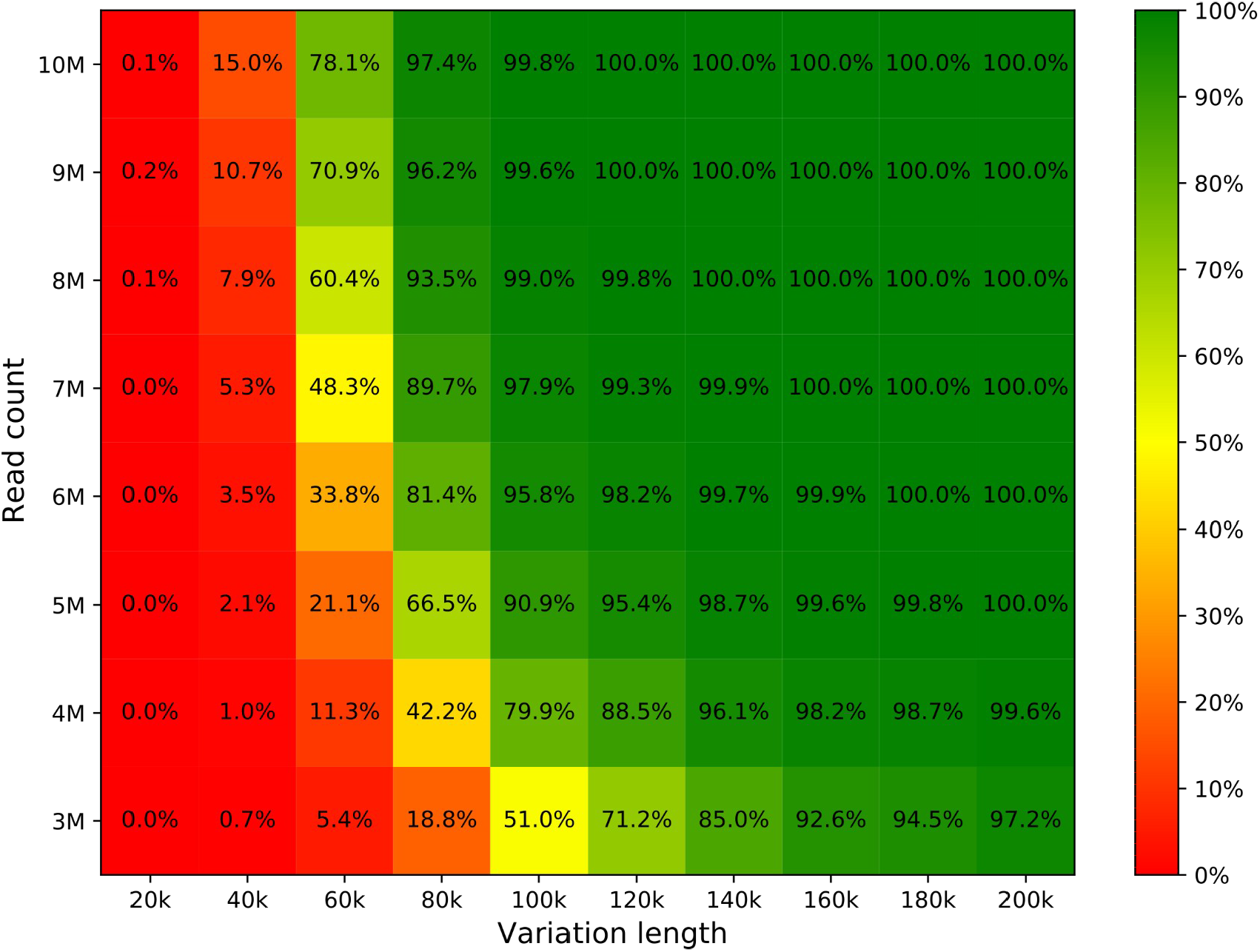
Prediction accuracy computed with *in-silico* analysis based on the length of variation and read count. Each cell number is generated from 8,300 simulations (100 randomly generated aberrations, 83 samples).

### 3.3 Validation of clinical samples

Finally, we ran an evaluation of samples analyzed previously in diagnostic settings using the aCGH method (Human Genome CGH Microarray 4×44K Agilent [28]) and GenomeScreen. The chosen aCGH method has 42,494 probes, which result in mean accuracy of detection of approximately 200 kb; however, the probes are focused mainly in gene regions and very sparsely in intergenomic regions, therefore the accuracy will be better within and worse outside of genes.

From the 106 tested samples, 58 did not show any detection on aCGH, and the rest contained 59 detections together (lengths from 39 kb to 146 Mb), from which GenomeScreen also detected all. The detections on GenomeScreen and on aCGH have excellent concordance – median 94.37% overlap (more data in Supplementary material Table 1). GenomeScreen furthermore detected 134 additional variations with ranges from 80 kb up to 1.48 Mb, mainly in regions with a low number of aCGH probes and protein coding genes, where aCGH has low coverage (Figure 4. and Supplementary material Table 1. and Figure 1.).

**Figure 4.**
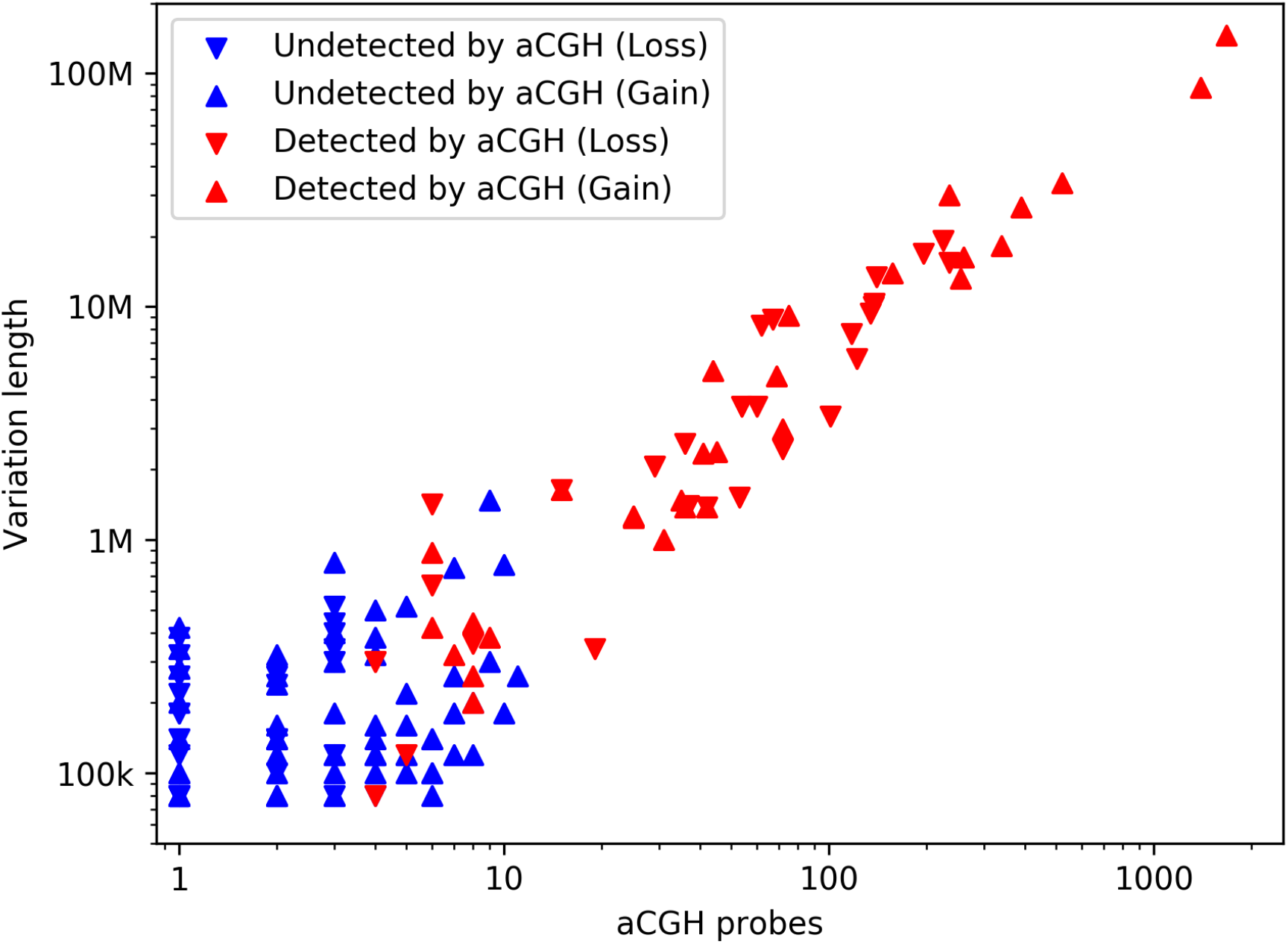
Detection of GenomeScreen (all) and aCGH (red) based on the variation length and number of aCGH probes in the detected interval (by GenomeScreen). Deletions and duplications are visualized by lower and upper triangles, respectively.

## 4. Discussion

GenomeScreen test is a result of evolving laboratory methods and bioinformatic tools validated in our laboratory and is currently available commercially. The genesis of the assay has begun with a basic NIPT test focused on noninvasive prenatal screening for three most common trisomies, continued with the addition of sex chromosomes aneuploidies, five selected microdeletions detection, and most recently moved to whole-genome scan for chromosomal microaberrations [18,24,25]. The common link between all these tests is the method based on low-coverage whole-genome sequencing. Because all the versions of mentioned NIPT tests are intended only for screening, we wanted to validate the method also for diagnostic purposes with much broader applicability in prenatal and postnatal diagnostics. One of the key applications is the replacement of aCGH as the confirmatory method in non-invasive prenatal diagnostics. Therefore, the method was in the pilot phase validated on plasma and amniotic fluid samples, later the analysis was extended to chorionic villi, placental tissue, blood, buffy coat, and fetal tissue.

GenomeScreen uses a binning approach and thus the genomic coordinates of detected variations are reported as a multiplier of the bin-size (20 kb) and thus it is not suitable for precise CNV detection at the level of exons. On the other hand, the aCGH method uses probes, which can be seen as variable size bins, where the resolution is equal to the probe distance (which is sometimes larger than the 20 kb bin-size). The precision of both the GenomeScreen and aCGH can be easily increased (by decreasing bin size and deeper sequencing in case of GenomeScreen, or by introducing new probes in case of aCGH), but these adjustments inevitably bring higher production cost.

The overall accuracy depends highly on the depth of sequencing (see Figure 3.). If we set the sequencing depth to achieve slightly favorable accuracy to the aCGH, the cost per sample is 2-3x lower for GenomeScreen. Furthermore, the turnaround time from submission of a sample to completion of the whole process including analysis takes less time in the case of GenomeScreen, usually 2 to 5 days, whereas the aCGH process may take up to 2 weeks when cultivation is required. The cultivation or DNA amplification is usually required in NIPT settings, since the amount of retrieved DNA is not sufficient for direct use of aCGH. However, even without these previous preparations, the hybridization process itself takes at least 3 days to deliver the result.

The disadvantage of GenomeScreen is the necessity to train the used normalization on at least 100 non-aberrated samples (the training on less samples results in filtration of unnecessarily numerous bins due to high variability), but we recommend using as much sample as possible for training. The training should be done separately for each sample type (and/or different laboratory protocol), however, the trained parameters are quite close for different sample types that we studied, and thus the parameters can be reused with only a slight decrease of accuracy and noise in CNV profiles. We did not experiment with different laboratory protocols, thus we can not assess how it can affect the training parameters. The need for re-training for different laboratory processing of the samples and/or sample types makes this approach difficult to test on datasets other than our own since the datasets available usually do not contain enough samples and information to train and test GenomeScreen. The study is based on analyses of 789 training and 106 control samples, with both groups of plasma type.

The false-positive rate of GenomeScreen is not studied in this work and should be adequately addressed in the future. However, the loss or gain of the (non-mosaic) deviation with a length of at least 100 kb is so substantial, that we do not expect to see any false positive detections.

One substantial, albeit only technological, advantage of the GenomeScreen method is the use of the same laboratories, protocols, chemistries, instruments, and laboratory technicians for both the screening NIPT test and also for the confirmatory GenomeScreen test. This was not possible in the case of the confirmatory aCGH test due to entirely different protocols and corresponding infrastructure and chemistry. The ability to use a method and its modifications with the same technical specification for screening as well as (subsequent and/or confirmatory) diagnostics is not often seen in laboratory medicine. Therefore, the presented study results fit into the trend of unification of processes on the side of laboratory work as well as bioinformatics and its utilization in different fields of clinical testing.

## 5. Conclusions

In this article, we presented a new method for CNV detection based on low-coverage whole-genome sequencing – GenomeScreen. We estimated its theoretical sensitivity and conducted a series of in-silico tests to estimate it in a semi-real setting. Afterward, we compared this method directly with a commonly used aCGH method on 48 control samples with known aberrations. The new method found all of the aberrations and even more aberrations mainly in intergenic regions, where the studied aCGH has poor coverage.

According to the presented results, GenomeScreen is currently able to detect almost all variations longer than 100 kb in mappable regions on the human genome. Moreover, it is cheaper and has faster turnaround times than the studied aCGH method. Thus, in presented laboratory settings, it is a favorable replacement for the more conventional aCGH method for detection of CNVs longer than to 100 kb.

## Data Availability

Data and scripts (Python 3.7) are available on the website https://github.com/marcelTBI/GenomeScreen .

https://github.com/marcelTBI/GenomeScreen

## Supplementary Materials

Supplementary materials can be found at a Google Drive folder https://drive.google.com/drive/folders/1VQU_Ijq5m0CdNzantB-tgG-3JmS79Id9.

## Author Contributions

Conceptualization, T.S., G.M., and J.B.; methodology, M.K, M.H., and G.M.; software, M.K.; validation, M.K., M.H., and G.M.; investigation, M.K.; resources, M.H.; data curation, M.K., M.H.; writing—original draft preparation, M.K.; writing—review and editing, M.K., M.H, G.M., and J.B..; visualization, M.K.; supervision, J.B.; funding acquisition, G.M., and T.S. All authors have read and agreed to the published version of the manuscript.

## Acknowledgments

We thank Ondrej Pös and Zuzana Kubiritová for their valuable comments and help with editing the manuscript text.

## Conflicts of Interest

We declare a potential competing financial interest in the form of employee contracts (see affiliations for each author) with Geneton Ltd. and TrisomyTest Ltd. Geneton Ltd. participated in the development of a commercial NIPT test in Slovakia; however, it is not a provider of this commercial test, but continues to do basic and applied research in the field of NIPT. On the other hand, TrisomyTest Ltd. is the commercial provider of NIPT testing in Slovakia. Its participation in the study was limited to the routine NIPT testing that generated the genomic results reused in our research. Related to this work, there are no patents, products in development, or marketed products to declare. The authors declare no other conflict of interest.

### Abbreviations

aCGH: array-based comparative genomic hybridization
CBS: circular binary segmentation
CNV: copy number variant
NGS: next-generation sequencing
NIPT: non-invasive prenatal testing
WGS: whole-genome sequencing

