## Supplementary figures and images for "Copy number variant detection with low-coverage whole-genome sequencing is a viable alternative to the traditional array-CGH"

### Supplemental Figure 1.

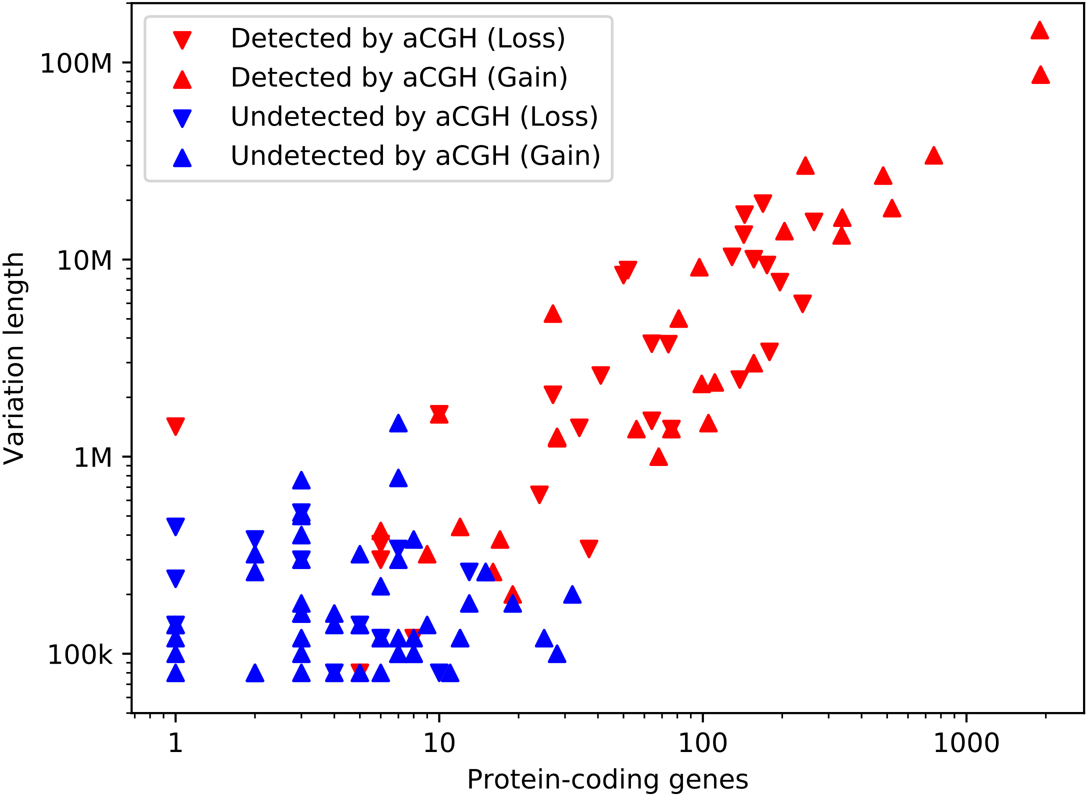
